# Development of the European Association for the Study of Obesity (EASO) Grade-Based Framework on the Pharmacological Treatment of Obesity: Design and Methodological Aspects

**DOI:** 10.1101/2025.04.07.25325370

**Authors:** Barbara McGowann, Andreea Ciudin, Jennifer L. Baker, Luca Busetto, Dror Dicker, Gema Frühbeck, Gijs H. Goossens, Matteo Monami, Benedetta Ragghianti, Paolo Sbraccia, Borja Martinez-Tellez, Euan Woodward, Volkan Yumuk

**Author notes:** BM and AC contributed equally. Corresponding authors: Barbara McGowan, Andreea Ciudin.

## Abstract

**Aim:** To describe the design and methodological aspects of the upcoming European Association for the Study of Obesity (EASO) Framework for the Pharmacological Treatment of Obesity utilizing currently available evidence.

**Methods:** An expert panel of 13 members, selected by EASO, developed the framework using the GRADE methodology to ensure transparent, evidence-based guideline development. Clinical questions were formulated using the Population, Intervention, Comparator, Outcomes (PICO) framework, focusing on the effectiveness and safety of European Medicines Agency-approved obesity management medications (OMM), including orlistat, naltrexone/bupropion, liraglutide, semaglutide, and tirzepatide. A comprehensive literature search will be conducted using Medline and Embase, including randomized controlled trials with a minimum duration of 48 weeks. Meta-analyses and network meta-analyses will be conducted to compare treatment effectiveness and safety profiles across various patient subgroups.

**Results:** The guidelines will target adults with a body mass index (BMI) ≥27 kg/m^2^ and at least one weight-related comorbidity or a BMI ≥30 kg/m^2^. The primary endpoint will be total body weight loss (TBWL%). Secondary outcomes will be changes in body composition (i.e., fat mass, fat-free mass), metabolic improvements (i.e., glucose levels, HbA1c, lipid profile), remission of obesity-related comorbidities (i.e., type 2 diabetes, obstructive sleep apnoea syndrome, metabolic dysfunction-associated steatotic liver disease, cardiovascular disease, and knee osteoarthritis), and improvements in mental health and quality of life. Preliminary analyses suggest that this framework will provide structured, individualized treatment recommendations based on the latest evidence.

**Conclusions:** The EASO framework aims to optimize pharmacological treatment for obesity through an individualized, evidence-based approach. By integrating clinical efficacy, safety outcomes, and patient-specific factors, these guidelines will support healthcare professionals in improving obesity management and its related comorbidities.

## 1. INTRODUCTION

Obesity is an adiposity-based chronic disease (ABCD)^1^ that significantly affects both physical and mental health^2-4^. Excessive and/or dysfunctional adipose tissue has detrimental effects on health, quality of life and life expectancy. Furthermore, significant complications associated with obesity include type 2 diabetes (T2D), ischaemic heart disease, heart failure, metabolic dysfunction-associated steatotic liver disease (MASLD), obstructive sleep apnoea syndrome (OSAS), knee osteoarthritis (KOA), mental disorders and certain forms of cancer, among others.

Current methods of obesity treatment include lifestyle intervention, endoscopic or surgical procedures and obesity management medications (OMM)^1^. However, most of these tools are often characterized by limited long-term adherence and efficacy or few/no available data on their effectiveness and safety^5^. In Europe, there are several European Medicines Agency (EMA)-approved OMM such as orlistat, naltrexone/bupropion, liraglutide, semaglutide, and tirzepatide. Additionally, in some countries, phentermine-topiramate is approved. The OMM, especially the analogues of peptides stimulated by nutrients, act at several levels in both the brain and peripheral tissues. Their primary effects include reducing appetite, enhancing energy metabolism —leading to weight loss, changes in body composition and positively influencing obesity-related complications.

The 2024 European Association for the Study of Obesity (EASO) framework for the diagnosis, staging and management of obesity in adults recommends that the treatment of obesity should aim at an individualised approach, reducing adiposity, improving adipose tissue function, managing obesity-related complications and, overall, improving the quality of life of patients with obesity. The present manuscript describes the design and methodological aspects of the upcoming EASO Framework for the Pharmacological Treatment of Obesity.

## 2. METHODS

### 2.1 Characteristics of the panel of experts

Panel members (n=13) identified by the EASO (Table S1), elected two coordinators according to EASO Guidelines (*BG and AC, Co-chairs of the Obesity Management Working Group of EASO*) and nominated the members of the Evidence Review Team (ERT). The ERT collected and analysed all available evidence, without participating in the definition of clinical questions, outcomes, and recommendations. The complete list of the panel and ERT members, including their roles, is reported in **Table S1**. All members reported a declaration of potential conflicts of interest (COI), which were collectively discussed to determine their relevance. The COIs were considered not relevant for this work. The panel of experts agreed to use the Grading of Recommendations, Assessment, Development, and Evaluation (GRADE) as described below and to formulate recommendations exclusively on results of meta-analyses of either placebo-controlled randomised clinical trials (RCT) or head-to-head RCT. The decision to exclude non-RCTs was made due to methodological concerns with observational studies (i.e. selection bias, prescription bias, etc.) and to comply with the Cochrane Manual, which recommends avoiding the use of non-randomised studies in systematic reviews and meta-analyses, particularly when assessing pharmacological interventions (https://training.cochrane.org/handbook/current/chapter-24).

### 2.2 GRADE methodology for the development of guidelines

The GRADE methodology is the most commonly adopted tool for transparently developing healthcare guidelines (https://training.cochrane.org/grade-approach). The formulations of any recommendation should be based on the best available evidence, which should be synthesised to achieve a reliable balance between benefits and harms, patient values, and preferences. Following GRADE methodology, the first step was to formulate clinical questions using the Population, Intervention, Comparator, Outcomes (PICO) framework (https://training.cochrane.org/grade-approach), as reported in **Table 1**. The panel identified several outcomes classified as critical, important, or less important after internal discussion. For critical outcomes, the ERT will perform formal meta-analyses, including all relevant trials fulfilling predefined search strategies and inclusion criteria, as reported in the results section.

**Table 1.**
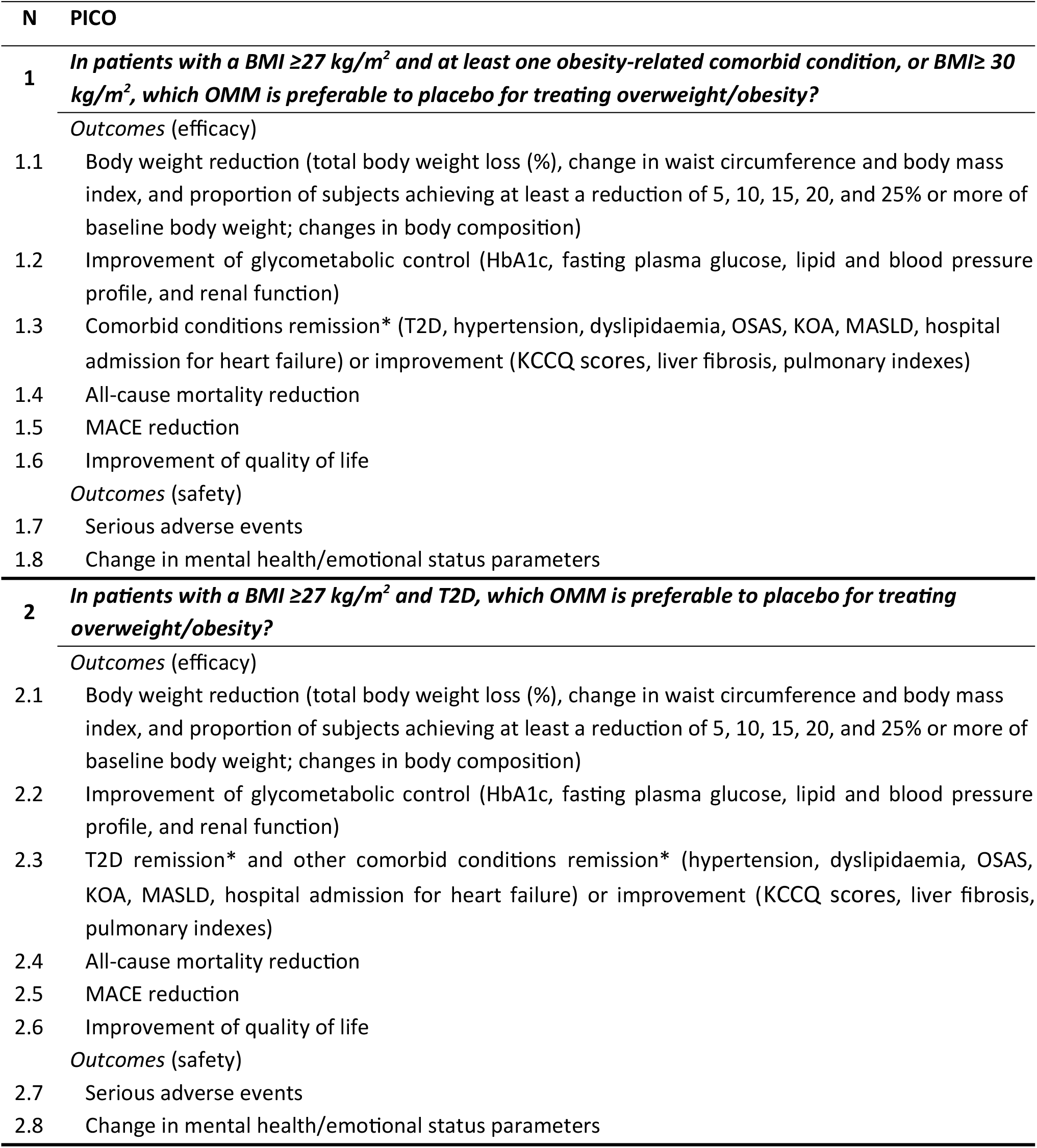
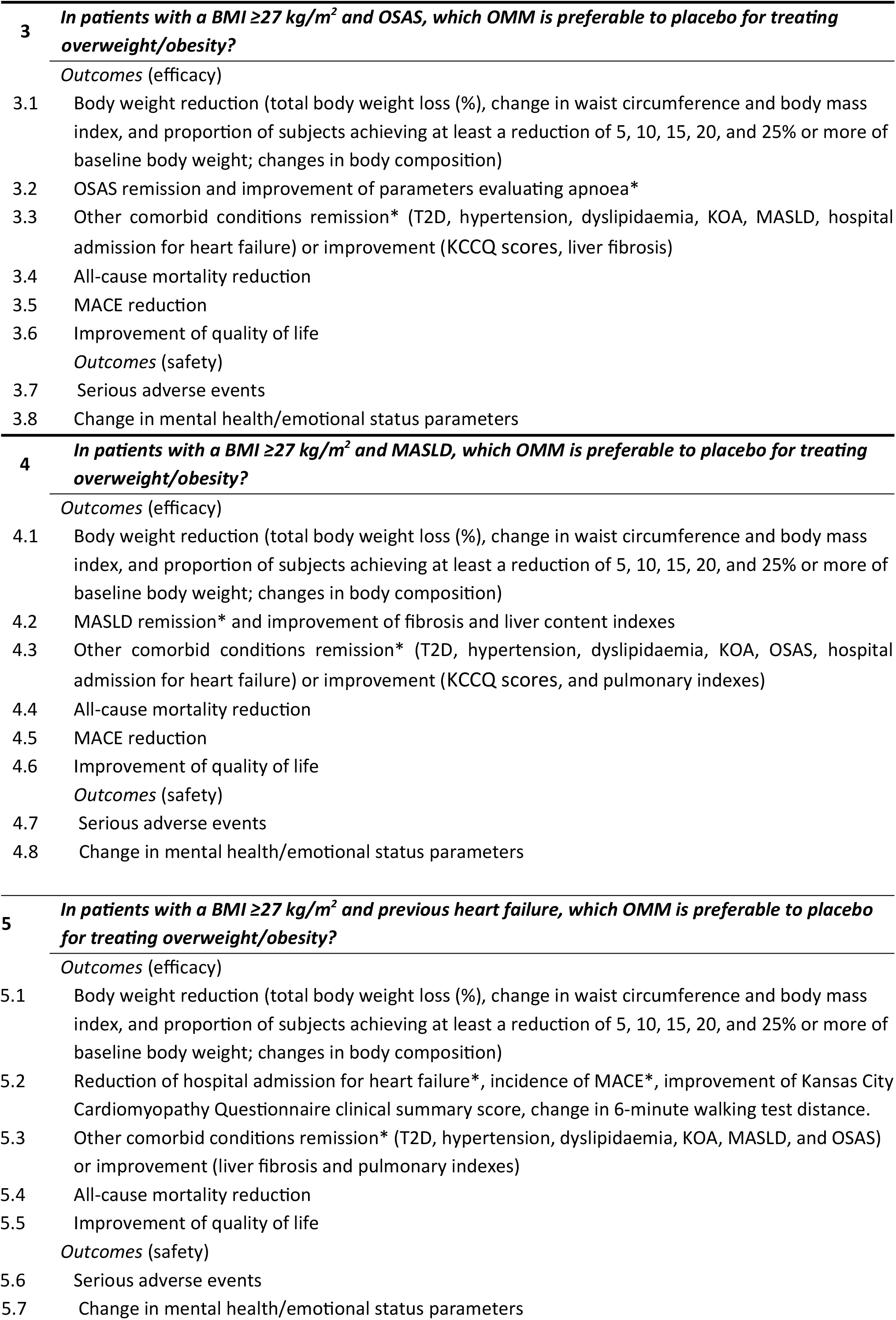
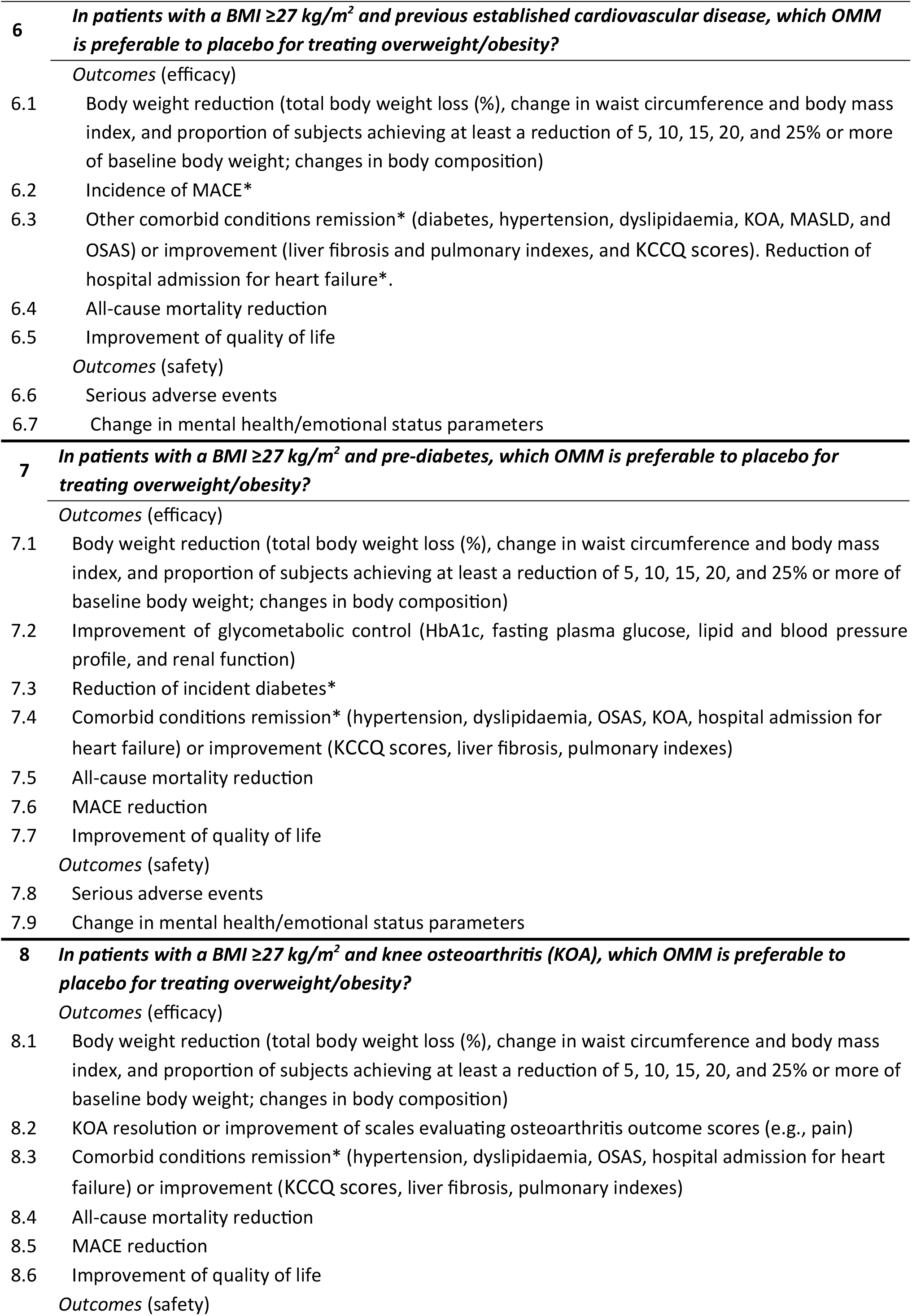
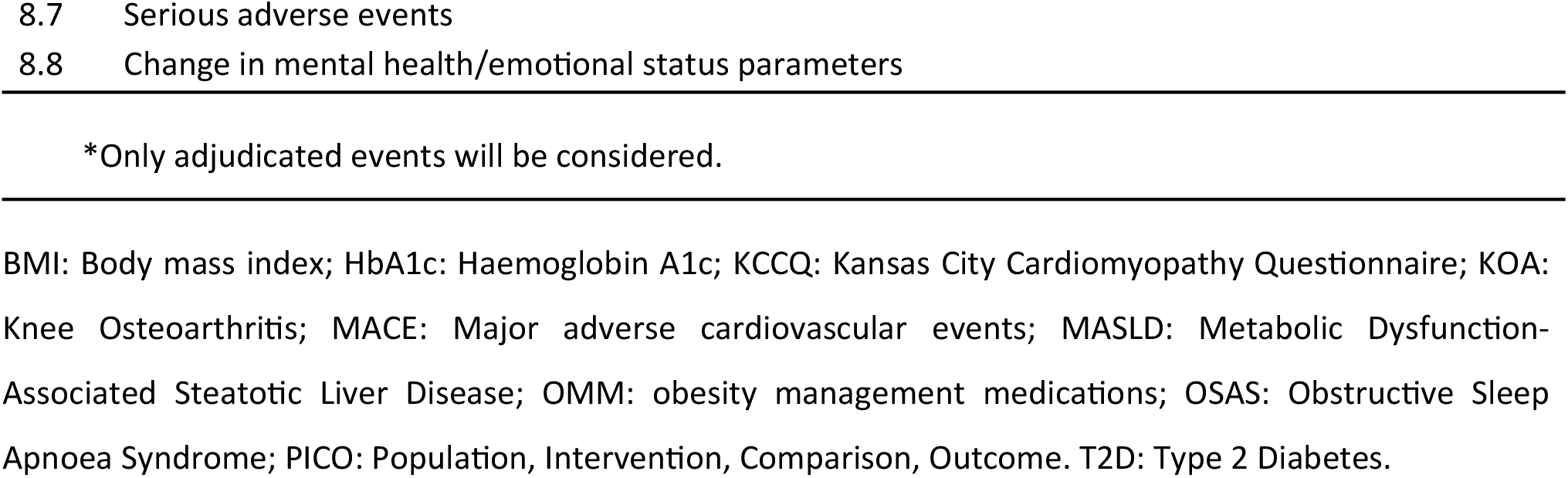
Population, Intervention, Comparator, Outcomes (PICO) definitions and related outcomes.

A crucial component of the GRADE methodology consists of evaluating the quality of evidence. This assessment considers the impact of several possible biases on the overall quality of the included studies. The domains considered will include: 1) evaluation of the methodological quality of the studies; 2) inconsistency (i.e., assessing the variability in study results); 3) indirectness (i.e., applicability of the evidence to the clinical question; 4) imprecision (i.e., evaluation of the certainty of the evidence; 5) publication bias (i.e., checking for selective publication of studies)^7^. The quality of evidence will be rated as high, moderate, low, or very low. This rating will be the basis for deciding the strength of each recommendation^8^ (i.e., strong or weak). Strong recommendations indicate high confidence in the benefits outweighing the risks; on the contrary, weak recommendations suggest that the benefits and risks are closely balanced or uncertain^8^. GRADE methodology will be used to assess the quality of the body of retrieved evidence. GRADEpro GDT software (GRADEpro Guideline Development Tool, McMaster University, 2015) will be used.

## RESULTS

This guideline will apply to adult patients either with a body mass index (BMI) ≥27 kg/m ^2^ and at least one weight-related comorbid condition or with BMI ≥30 kg/m^2^. In developing these guidelines, healthcare systems, infrastructures and human and financial resources across European regions will be considered.

### Panel of experts

The present guidelines will be developed by a multidisciplinary team composed of specialists in endocrinology and nutrition, internal medicine, epidemiology, and basic and clinical scientists (**Table S1**). The panel proposed eight clinical questions, which were approved without the need for further discussion (Table 1). The approved questions and their related approved critical outcomes are reported in Table 1. For critical PICO, formal systematic reviews and network meta-analyses will be performed to rank the effectiveness and safety of OMM considered in the present guidelines.

### Search strategy

A Medline and Embase search will be performed without establishing a starting date, up to 31^st^ January 2025, using the following keywords: obesity AND (orlistat OR naltrexone OR bupropion OR topiramate OR phentermine OR liraglutide OR semaglutide OR tirzepatide). The name of the OMM is placed in chronological order, which means according to when these drugs were available in the market. We will include only RCTs enrolling patients either with BMI ≥27 kg/m^2^ and at least one weight-related comorbid condition or obesity (BMI ≥30 kg/m^2^) and comparing different OMM either versus placebo/no therapy (add-on to lifestyle intervention) or active comparators, with a duration of at least 48 weeks.

### Interventions included for guideline development

The interventions that were considered to develop the guideline included OMM (i.e., orlistat 360 mg/day, naltrexone SR/bupropion 32-360 mg/day, topiramate/phentermine 15-92 mg/day, liraglutide 3.0 mg/day, semaglutide 2.4 mg/week, tirzepatide 10-15 mg/week versus placebo/none), lifestyle interventions or active comparators.

### Endpoints

The principal endpoint will be Total Body Weight Loss (TBWL%) at the study endpoint in clinical studies designed primarily to investigate the effects of OMM on patients for whom the major issue is body weight reduction and not the control of other obesity-associated medical conditions. For patients included in the other subgroups (e.g., T2DM, prediabetes, previous heart failure, etc.) the primary endpoint is reported in Table S2.

Secondary endpoints (for all subgroups) will include the following (placebo-subtracted absolute differences from baseline, if not otherwise specified):

a. TBWL% at 52 weeks, 53-104 weeks, and ≥105 weeks
b. Change in endpoint BMI and waist circumference
c. Change in Fat Mass (FM), Subcutaneous Fat (SFM) and Visceral Fat (VFM)
d. Change in Fat-Free Mass (FFM)
e. Proportion of subjects achieving at least 5, 10, 15, 20, and 25% of body weight reduction (odds ratio)
f. T2D (defined as the achievement of HbA1c<6.5%), hypertension (defined as the cessation of anti-hypertensive drugs), dyslipidaemia (defined as the cessation of anti-hypertensive drugs), MASH (defined as no steatotic liver disease or simple steatosis without steatohepatitis), reduction of at least one stage of liver fibrosis, OSAS remission (defined as Apnoea-Hypopnea Index (AHI) <5 or AHI of 5 to 14), and reduction of hospital admission for Heart Failure (odds ratio)
g. Serious Adverse Events (SAE), all-cause mortality, or MACE (odds ratio)
h. Fasting Plasma Glucose, Glycated Haemoglobin (HbA1c), lipid profile, blood pressure, eGFR, creatinine, and albuminuria
i. Improvement of Kansas City Cardiomyopathy Questionnaire scores, liver fibrosis and fat content, and pulmonary indexes.
j. Change in mental health parameters
k. Endpoint Quality of Life (QoL)

### Statistical analyses

To compare and rank the efficacy and safety of OMMs, depending on the data available, we will perform NMA, pairwise meta-analyses or subgroup meta-analyses. Pre-planned analyses will be performed based on the following patient baseline characteristics as specified in Table 1. The endpoints considered critical (primary and secondary) for each subpopulation of patients are specified in Table S2.

Treatment effects will be reported as weighted mean differences (WMD) with 95% confidence intervals (CI) for continuous outcomes and odds ratios (OR) with 95% CI for categorical outcomes. Placebo will serve as the reference category for ranking treatments. In all analyses between-study heterogeneity will be quantified using I^2^ statistics. If substantial heterogeneity is detected (I^2^ >50%), we will explore potential sources through subgroup analyses whenever possible.

We decided to use NMA as the principal methodology, due to its capability of allowing indirect comparisons when direct head-to-head trials are unavailable and because it combines direct and indirect evidence to provide a comprehensive estimate of treatment effects. We will use a random-effects model within a generalised pairwise modelling (GPM) framework. Before beginning the NMA, we will evaluate transitivity by examining key effect modifiers across treatment comparisons. Studies with significant differences in effect modifiers will be considered in sensitivity analyses. These analyses will be performed in in MetaXL (www.epigear.com).

If a NMA is not feasible due to less than 10 trials, disconnected networks, or clinical heterogeneity, we will conduct pairwise meta-analyses where direct comparisons are available. We will additionally assess statistical heterogeneity using Cochran’s Q test (p < 0.10 indicating heterogeneity). If heterogeneity is high, we will explore sources using subgroup and sensitivity analyses.

If pairwise meta-analyses are not feasible due to due to a lack of direct evidence or high heterogeneity, we will perform separate meta-analyses for predefined subgroups (e.g., OMM vs. placebo, OMM vs. LSI). These analyses will use random effects models and be performed in RevMan, Version 5.3 (Copenhagen: The Nordic Cochrane Centre, The Cochrane Collaboration, 2014).

If statistical pooling is not possible, we will provide a qualitative synthesis of the findings. We will use the Risk of bias assessment tool to judge the reliability of the study. We will provide descriptive comparisons of treatment effects across studies based on the study population, OMM, comparison group, and outcome similarities.

We will assess small-study effects and publication bias using funnel plots (for outcomes with ≥10 studies), Egger’s test (for continuous outcomes), and Harbord’s test (for binary outcomes).

## DISCUSSION

While the history of obesity pharmacotherapy has been fraught with setbacks, safe and effective OMMs are currently available. However, so far, no specific recommendations have been developed. Thus, the areas covered by the clinical questions proposed by the expert panel include indications for the appropriate use of OMM in different subgroups of patients. Several OMM have been assessed in people living with obesity and obesity-related medical conditions^10,11^, such as MASLD, OSAS, Heart failure, MACE, and T2D. These upcoming guidelines proposed by EASO will provide an algorithm to tailor the use of OMM by analysing available evidence for several categories of individuals.

The decision to adopt a certain pharmacological treatment will be determined in some cases by its intrinsic efficacy to reduce TBWL, and in other cases it will be driven by the effect of the OMM to improve specific complications related to obesity, aligning the OMM recommendations with the EASO framework for the diagnosis, staging and management of obesity as an ABCD^1,12^. Based on the definition of obesity as an ABCD, some patients may be living with fat-mass disease, for example, patients with obesity and OSAS, where the goal would be to induce TBWL. Others may be affected by “sick-fat disease”, and their goal would be the management and improvement of obesity-related medical conditions, in addition to TBWL. Moreover, the choice of a specific therapeutic option should be based on an accurate assessment of the risk-benefit ratio. Therefore, safety outcomes have been included for all proposed PICO, concurring with the development of recommendations.

Transparency in developing a GRADE-based guideline is one of the major determinants of its quality^13^. The GRADE manual recommends the publication of clinical questions, relevant outcomes, and summaries of evidence for each outcome^14^. The panel of experts involved in the present project decided to go beyond these requirements, pre-emptively publishing in extenso the entire process leading to clinical questions and definition of critical outcomes. In addition, the search strategy and inclusion criteria for the systematic review and meta-analysis for each outcome has been reported in the present study, allowing a transparent reproducibility of the whole process. Notably, the panel decided to extensively publish in peer-reviewed journals all systematic reviews and meta-analyses needed for formulating these guidelines.

## Data Availability

All data produced in the present study are available upon reasonable request to the authors

## Statement of Ethics

Not applicable. This article does not contain any studies with human participants or animals performed by any of the authors.

## Conflict of Interest Statement

Barbara McGowan (BMG) has received speaker and/or advisory fees from Novo Nordisk, Eli-Lilly, Astra Zeneca, Janssen, Pfizer, MSD and a research grant from Novo Nordisk. BM is a shareholder of Reset Health.

Andreea Ciudin (AC) has received speaking fees from Astra Zeneca, Boehringer-Ingelheim, Eli-Lilly, Novo Nordisk, Sanofi, Menarini and research grants from Eli Lilly, NovoNordisk and Menarini. Member of the DMC of Boehringer Ingelheim.

Jennifer L. Baker (JLB) has received a consulting fee and is an advisory board member for Novo Nordisk A/S with fees paid to her institution.

Luca Busetto (LB) has received payment of honoraria from Eli Lilly, Novo Nordisk, Boehringer Ingelheim, Pfizer, Bruno Farmaceutici, Regeneron, Rythm Pharmaceuticals and Pronokal as speaker and/or member of advisory boards.

Dror Dicker (DD) has received speaker and advisory board fees from Boehringer-Ingelheim, Eli-Lilly, Novo Nordisk, Astra Zeneca and research grants from Eli Lilly, Novo Nordisk and Boehringer Ingelheim.

Gema Frühbeck (GF) has received payment of honoraria from Eli Lilly, Novo Nordisk, Regeneron and Astra Zeneca as speaker and/or member of advisory boards, and payment of honoraria as member of the OPEN Spain Initiative.

Gijs H. Goossens (GHG) has no relevant conflicts of interest to declare related to this article. Matteo Monami (MM) has received speaking fees from Astra Zeneca, Bristol Myers Squibb, Boehringer-Ingelheim, Eli-Lilly, Merck, Novo Nordisk, Sanofi, and Novartis and research grants from Bristol Myers Squibb.

Benedetta Ragghianti (BR) has no conflicts of interest to declare.

Paolo Sbraccia (PS) received payment of honoraria and consulting fees from Boehringer Ingelheim, Chiesi, Novo Nordisk, Eli Lilly, Pfizer, and Roche as a member of advisory boards.

Borja Martinez-Tellez (BMT) has received grants from the EASO New Clinical Investigator Award 2024 and the EFSD Rising Star 2024, both supported by the Novo Nordisk Foundation.

Euan Woodward (EW) has no conflicts of interest to declare.

Volkan Yumuk (VY) was engaged in advisory boards and lectures with: Novo Nordisk, Eli Lilly, Rhythm, Regeneron.

## Funding Sources

Not applicable

## Author Contributions

All the authors approved the final version of this manuscript. Dr. Barbara McGowan and Andreea Ciudin are the persons who take full responsibility for the work as a whole, including the study design, and the decision to submit and publish the manuscript.

Authors involvement in each of the following points:

1. Design: BMG, AC, JLB, LB, DD, GF, GHG, MM, PS, EW, VY
2. Data collection: MM, BR, BMT
3. Analysis: MM, BR
4. Writing the manuscript: BMG, AC, JLB, DD, GHG, MM, PS, BMT
5. Review of final draft: all the authors

## Data Availability Statement

All data generated or analyzed during this study are included in this article [and its supplementary material files]. Further enquiries can be directed to the corresponding author.

## Supplementary material

**Table S1.** Characteristics and tasks of all panelists.

| **N** | **Name** | **Profession** | **Role** |
| --- | --- | --- | --- |
| 1 | Barbara McGowen | Endocrinologist | Coordinator |
| 2 | Andreea Ciudin | Endocrinologist | Coordinator |
| 3 | Jennifer L. Baker | Epidemiologist | Panelist |
| 4 | Luca Busetto | Internist | Panelist |
| 5 | Dror Dicker | Internist | Panelist |
| 6 | Gema Frühbeck | Clinical Scientist | Panelist |
| 7 | Gijs H. Goossens | Biologist | Panelist |
| 8 | Matteo Monami | Diabetologist | Methodologist |
| 9 | Benedetta Ragghianti | Endocrinologist | ERT |
| 10 | Paolo Sbraccia | Internist | Panelist |
| 11 | Borja Martinez-Tellez | Human physiologist | ERT |
| 12 | Euan Woodward | Executive director<br>EASO | Management |
| 13 | Volkan Yumuk | Endocrinologist | Panelist |

ERT: Evidence Review Team

**Table S2.** Outcomes considered critical (primary and secondary) by the panel of experts for each subpopulation of patients with overweight/obesity.

|  |
| --- |
| **Pre-existing comorbid condition** |
| **Type 2 diabetes mellitus** |
| *Primary endpoint:* complete diabetes remission* |
| *Other critical endpoints:* Body weight reduction (total body weight loss % - TBWL%); lipid and blood pressure profile, and renal function), and improvement of metabolic control (glycosylated hemoglobin - HbA1c and fasting plasma glucose - FPG). |
| **Pre-diabetes** |
| *Primary endpoint:* normoglycemia restoration* |
| *Other critical endpoints:* Body weight reduction (total body weight loss % - TBWL%); lipid and blood pressure profile, and renal function), reduction of incident diabetes*, and improvement of metabolic control (glycosylated hemoglobin - HbA1c and fasting plasma glucose - FPG). |
| **Established cardiovascular disease** |
| *Primary endpoint:* Incidence reduction of Major Adverse Cardiovascular Events (MACE)* |
| *Other critical endpoints:* Body weight reduction (total body weight loss % - TBWL%), all-cause and cardiovascular mortality reduction. |
| **Heart failure** |
| *Primary endpoint:* Reduction of hospital admission for heart failure* |
| *Other critical endpoints:* Body weight reduction (total body weight loss % - TBWL%), incidence of Major Adverse Cardiovascular Events (MACE)*, improvement of Kansas City Cardiomyopathy Questionnaire clinical summary score, change in 6-minute walking test (6-m WT) distance. All-cause and cardiovascular mortality reduction |
| **Obstructive sleep apnea syndrome** |
| *Primary endpoint:* OSAS remission * |
| *Other critical endpoints:* Body weight reduction (total body weight loss % - TBWL%) and improvement of parameters evaluating apnea* |
| **Metabolic dysfunction-associated steatotic liver disease** |
| *Primary endpoint:* MASH remission* |
| *Other critical endpoints:* Body weight reduction (total body weight loss % - TBWL%) and improvement of fibrosis and liver indexes. |
| **Knee osteoarthritis** |
| *Primary endpoint:* KOA improvement assessed with scales evaluating osteoarthritis outcome scores (Western Ontario and McMaster Universities Osteoarthritis Index – pain and physical-function score), |
| *Other critical endpoints:* Body weight reduction (total body weight loss % - TBWL%), improvement of 6-m walking distance, and opioids use. |

\*Only adjudicated events will be considered. For PICO 1 all outcomes will be considered critical; for PICO 2- 8 only underlined outcomes will be analyzed separately in RCTs specifically designed for that subpopulation of patients with overweight/obesity.

## Notes

### Competing Interest Statement

The authors have declared no competing interest.

### Funding Statement

This study did not receive any funding

